# MASSIVE SCABIES OUTBREAK IN ROHINGYA REFUGEE CAMPS, COX’S BAZAR: SEASONALITY AND ASSOCIATION WITH CLIMATIC FACTORS (2021–2023)

**DOI:** 10.64898/2026.01.06.26343570

**Authors:** Charls Erik Halder, Md Abeed Hasan, James Charles Okello, Muhammad Feroz Khan, Emmanuel Roba Soma, Julekha Tabassum Poly, Hamim Tassdik, Md Sobuz Hosen, U Maung Prue, Md Abdul Hannan

## Abstract

**Background:** Scabies is a common skin condition and poses a substantial disease burden in resource-poor tropical settings. The Rohingya refugee camps in Cox’s Bazar, Bangladesh represent one of the world’s largest and most protracted humanitarian crises. Using three years of data from 2021 to 2023 and applying rigorous analytical methods, this study describes the seasonality of scabies and examines its association with climatic factors.

**Methodology:** This is a retrospective observational study conducted in the Rohingya refugee camps and adjacent host communities in Ukhiya and Teknaf, Cox’s Bazar. All patients clinically diagnosed with scabies and who received treatment at 35 International Organization for Migration (IOM)–supported health facilities between 1 January 2021 and 31 December 2023 were included. Climate data, including daily mean, minimum and maximum temperature and total and maximum rainfall, were obtained from the Bangladesh Meteorological Department. Seasonal–trend decomposition using Loess (STL) was applied. Associations between climatic variables and the decomposed seasonal component of scabies cases, as well as overall scabies case counts, were assessed using Pearson correlation tests.

**Results:** A total of 323,106 new scabies cases were reported from IOM-supported health facilities between January 2021 and December 2023. Children aged under 5 years and 6–18 years accounted for the highest proportion of cases (32.08% and 38.95%, respectively). The average monthly number of scabies cases was highest in November (12,625) and lowest in May (5,862). Case numbers increased from November to February (high season), with a peak between October and November, and declined between April and June (low season). An inverse relationship was observed between temperature and scabies incidence, with higher case numbers during cooler months and lower numbers during warmer months. Pearson correlation analysis demonstrated a strong and significant negative correlation between the seasonal component of scabies and maximum (r = −0.492, p = 0.002), minimum (r = −0.506, p = 0.002), and mean temperature (r = −0.525, p = 0.001). No significant association was observed between scabies seasonality and humidity or rainfall.

**Conclusion:** This study identified a distinct seasonal pattern of scabies, with higher caseloads during late autumn and winter (October to February) and lower caseloads during summer months (April to June). Temperature showed a strong negative association with the seasonal component of scabies. These findings may inform the timing of public health strategies, including mass drug administration, intensified case management, and social and behavioural change communication, in humanitarian settings.

## INTRODUCTION

Scabies, a common skin condition, shares a substantial disease burden in resource-poor tropical settings. Classified as one of the neglected tropical diseases, scabies affects approximately 400 million individuals annually (1) . It is a parasitic infestation caused by the mites, *Sarcoptes scabiei var hominis*, that burrow into the skin and lay eggs, causing intense itching and a rash. Scabies is highly contagious and transmitted through skin-to-skin contact. Complications of scabies include secondary bacterial skin infections and subsequent immune-mediated complications, specially, acute post-streptococcal glomerulonephritis and rheumatic heart disease (1–5).

While scabies occurs in almost every country globally, it is more prevalent in tropical countries where the climate is characteristically hot and humid (6,7). Disadvantaged people living in crowded and resource-poor setting are particularly vulnerable to scabies due to poor and overcrowded living conditions. Therefore, displaced populations and refugees living in camp settings are at disproportionately higher risk of scabies infestation and outbreak. Overcrowded living conditions, poor access to water, sanitation, and hygiene facilities, difficulty in cleaning and washing, limited access to treatment and healthcare, lack of medical supplies, stigma and discrimination exacerbate the risk of scabies outbreak in refugee camp settings (8,9).

The Rohingya refugee camps in Cox’s Bazar, Bangladesh is one of the world’s largest and most protracted humanitarian crises. Following decades of persecution and repeated displacements from Myanmar’s Rakhine State, the Rohingya community experienced a major influx into Bangladesh in August 2017. As of December 31, 2023, approximately 1,143,096 refugees are living in densely populated camps in Cox’s Bazar (10). The refugees as well as disadvantaged host communities surrounding the camps are at heightened risk of infectious disease outbreaks resulting from the overcrowded living conditions, fragile shelters, and insufficient water, sanitation, and hygiene (WASH) facilities. Frequent massive fire incidents, heavy monsoon, landslides and flashfloods further worsen the living conditions and vulnerabilities of the refugees and host communities.

Over the years, the camps have experienced outbreaks multiple infectious diseases, including diphtheria, measles, COVID-19, and acute watery diarrhea (AWD)/cholera (11–15). In our earlier study from this research series, we reported on the epidemiology of a large scabies outbreak in Rohingya refugee camps and host communities, with 384,852 cases from 2021 to 2024, where children and women had a disproportionate higher attack rate. Such persistent high prevalence of scabies places additional strain on the already overburdened healthcare system, increases pressure on the health economics in the resource poor setting and significantly impacts the quality of life of the community.

Many infectious diseases exhibit well established seasonal patterns, understanding which enables the public health practitioners and policy makers for timely placement of prevention and response measures. Seasonal patterns of scabies were documented in multiple studies (16–18). Reported seasonal patterns varied region to region. For instance, while studies in Taiwan reported higher prevalence of scabies in Winter, South Korea reported that in Autumn. Brazil, a tropical country, reported no seasonal variation (17–19). Bangladesh has a tropical monsoon climate characterized by distinctive monsoon, summer and winter. No studies have so far found that reported seasonality of Scabies in Bangladesh or any country that share similar climate in South Asia.

Globally, only a few studies analyzed the impact of climate factors, especially, temperature, rainfall and humidity, on the seasonality and trend of scabies (17,20). Since public health situation and human behavior have substantial impact on the transmission of scabies, it is theoretically difficult to distinguish the impact of climate factors on the scabies trend. However, impact of climate factors on scabies burden is essential to know for proper preparedness for and prevention of the scabies outbreak. Using the data of three years from 2021 to 2023 and applying rigorous scientific methods, this study describes the seasonality of scabies and examines its association with climatic factors. The findings of the research may assist health programmes and policymakers to timely plan public health measures to prevent and early respond to surge of scabies in the Rohingya refugee camps as well as similar resource poor and climatic settings.

## METHODOLOGY

### Study design

This is the second study under an umbrella of research on the epidemiology and economic impact of scabies in the Rohingya refugee camps. It is a retrospective observational study using de-identified and anonymized routine health data generated through standard clinical service delivery and public health surveillance activities implemented by the International Organization for Migration (IOM). This study is reported in accordance with the Strengthening the Reporting of Observational Studies in Epidemiology (STROBE) guidelines for observational studies.

### Study site and population

The study was conducted in the Rohingya refugee camps and adjacent host communities in Ukhiya and Teknaf, Cox’s Bazar, Bangladesh. All patients who were clinically diagnosed with scabies and received treatment and care at 35 IOM-supported health facilities between 1 January 2021 and 31 December 2023 were included. Data beyond 2023 were not included because mass drug administration (MDA) activities were implemented from 2024 onwards, which substantially altered scabies transmission patterns and could confound the assessment of seasonal trends examined in this study.

### Data collection

#### Scabies Data

De-identified and anonymized aggregated scabies case data were extracted from the IOM Cox’s Bazar Health Information Management System for the period January 2021 to December 2023. Data were originally collected using the IOM Cox’s Bazar outpatient department (OPD) reporting forms, entered electronically through Kobo Toolbox, and synchronized to a central database. Data quality checks, including verification and cleaning, were conducted routinely as part of standard surveillance procedures.

For this study, aggregated variables including age group, sex, nationality (host or refugee), and location were used to describe the demographic characteristics of scabies cases. Only day-wise counts of scabies cases were used for time-series and seasonal analyses. Individual-level identifiers were not collected, accessed, or available to the study team at any stage. The anonymized aggregated dataset was accessed for research purposes between 15 January 2024 and 30 March 2024.

All scabies cases were clinically diagnosed and managed at IOM-supported health facilities in accordance with Médecins Sans Frontières (MSF) clinical guidelines. Topical 5% permethrin cream was the primary treatment used for scabies management.

#### Climate Data

Climate data, including daily mean, minimum, and maximum temperature as well as total and maximum rainfall, were obtained from the Bangladesh Meteorological Department for the corresponding study period.

### Statistical analysis

The cleaned dataset was summarized using descriptive statistics to present the distribution of scabies cases by demographic characteristics, including age group, sex, location, and nationality. A combined day-wise dataset was created by merging daily scabies case counts with corresponding climate variables and subsequently aggregated to monthly values for seasonal analysis.

Monthly box plots were generated to visualize the distribution, central tendency (mean and median), and variability of scabies cases and to explore seasonal patterns. Seasonal–trend decomposition using Loess (STL) was applied to decompose the time series into trend, seasonal, and remainder components. Associations between climatic variables and both the decomposed seasonal component and overall scabies case counts were assessed using Pearson correlation tests. A p-value <0.05 was considered statistically significant.

Statistical analyses and visualizations were conducted using R (RStudio) and Microsoft Excel, including tabulation, graphical analyses, and regression-based procedures.

## ETHICAL CONSIDERATION

This study involved a secondary analysis of routinely collected public health surveillance data generated during scabies outbreak response activities in the Rohingya refugee camps and surrounding host communities in Cox’s Bazar, Bangladesh. Data were originally collected as part of ongoing clinical service delivery and outbreak monitoring, not for research purposes.

Ethical approval for the secondary analysis of anonymized and aggregated data was obtained from the Ethical Review Board of Cox’s Bazar Medical College Hospital (Approval No.: CoxMC/2023/017). Administrative clearance was also granted by the Office of the Civil Surgeon and the Refugee Relief and Repatriation Commissioner, the authorities responsible for oversight of health services in the Rohingya refugee camps.

The study used fully de-identified and aggregated data. Individual-level identifiers were not collected, accessed, or available to the study team at any stage. As the analysis was conducted using anonymized secondary data, the requirement for informed consent was waived by the ethics committee.

The anonymized aggregated dataset was accessed for research purposes between 15 January 2024 and 30 March 2024. The study was conducted in accordance with relevant national regulations and institutional guidelines governing the ethical use of routine public health data. This secondary analysis was classified as public health surveillance research and posed no more than minimal risk to participants.

## RESULTS

The study included a total of 323,106 new scabies cases reported from the IOM health facilities from January 2021 to December 2023. Table 1 demonstrates the demographic characteristics of the patients infected with scabies. Mean age of the patients was 14.6 and median age was 9. Children of both age group (below 5 years and 6 – 18 years) had the highest proportion of infestation (32.08% and 38.95% respectively). Three-quarters of the patients were from Ukhiya Upazila, and the rest were from Teknaf. More than 80% of the patients were Rohingya refugees, while the rest were from the surrounding host communities.

**Table 1.** Demographic characteristics of patients with scabies (2021 to 2023)

| Variable | Category | Scabies Count | Percentage (%) |
| --- | --- | --- | --- |
| **Overall** | Total | 323,106 | 100% |
|  | Mean age (years) |  | 14.6 |
|  | Median age (years) |  | 9 |
| **Sex** | Female | 171,164 | 52.97% |
|  | Male | 151,942 | 47.03% |
| **Age group** | 0 – 5 years | 103,656 | 32.08% |
|  | 6 – <18 years | 125,845 | 38.95% |
|  | 18 – <30 years | 43,830 | 13.57% |
|  | 30 – 60 years | 38,975 | 12.06% |
|  | 60 years+ | 10,800 | 3.34% |
| **Upazila** | Ukhiya | 245,148 | 75.87% |
|  | Teknaf | 77,958 | 24.13% |
| Community | Host | 59,959 | 18.56% |
|  | Refugee | 263,147 | 81.44% |

Figure 1 illustrates time series plots show daily values of average, minimum, and maximum temperature, relative humidity, total rainfall, and number of scabies cases. The visual demonstrates a temporal inverse association between temperature and scabies incidence.

**Figure 1.**
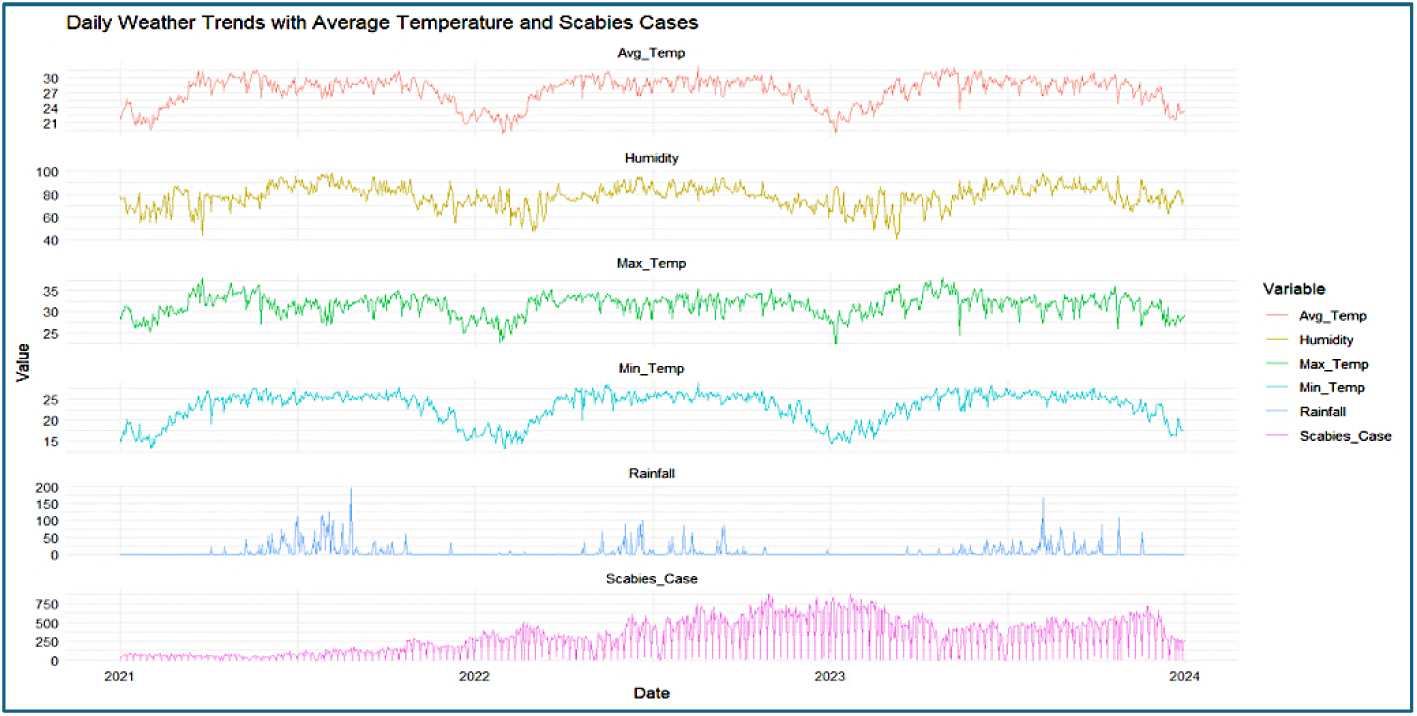
Daily weather and scabies trends from 2021 to 2023 in Cox’s Bazar refugee camps.

Table 2 summarizes the monthly climate variables and the number of scabies cases, averaged over three years from 2021 to 2023. The average temperature was highest in April (33.98°C) and lowest in January (16.57°C). Based on mean temperature, January to February were the coolest months, while April to May were the warmest months. The average total rainfall was highest in August (732 mm). Humidity was also highest in the same month (87.60%).

**Table 2.** Average monthly climate variables and number of scabies (2021 to 2023)

| Month | Temperature (°C) |  |  | Total<br>Rainfall<br>(mm) | Humidity<br>(%) | Scabies<br>Cases |
| --- | --- | --- | --- | --- | --- | --- |
|  | Max | Min | Mean |  |  |  |
| Jan | 27.97 | 16.57 | 22.27 | 2.67 | 70.00 | 9,698 |
| Feb | 29.47 | 17.49 | 23.48 | 10.00 | 68.50 | 8,704 |
| Mar | 32.90 | 22.47 | 27.68 | 11.33 | 70.28 | 7,697 |
| Apr | 33.98 | 25.17 | 29.58 | 50.33 | 75.07 | 6,299 |
| May | 33.39 | 25.71 | 29.55 | 185.33 | 78.37 | 5,862 |
| Jun | 31.50 | 25.51 | 28.51 | 597.33 | 86.59 | 7,764 |
| Jul | 31.83 | 25.84 | 28.83 | 505.00 | 85.22 | 7,948 |
| Aug | 31.43 | 25.56 | 28.50 | 732.00 | 87.60 | 10,126 |
| Sep | 32.23 | 25.65 | 28.94 | 297.33 | 85.22 | 9,215 |
| Oct | 32.84 | 25.09 | 28.96 | 204.67 | 81.92 | 11,667 |
| Nov | 31.94 | 22.55 | 27.25 | 31.67 | 73.10 | 12,625 |
| **Dec** | 29.14 | 18.94 | 24.17 | 19.67 | 73.14 | 10,096 |

The average number of scabies cases were highest in November (12,625) and lowest in May (5,862).

Figure 2 shows boxplot for monthly distribution of scabies cases from 2021 to 2023. October to February are the high-season months with higher monthly average numbers of reported scabies cases. October and November are the peak months with the highest means and medians. January and February months have high box heights, indicating variations in number of scabies cases in these months across the years.

**Figure 2.**
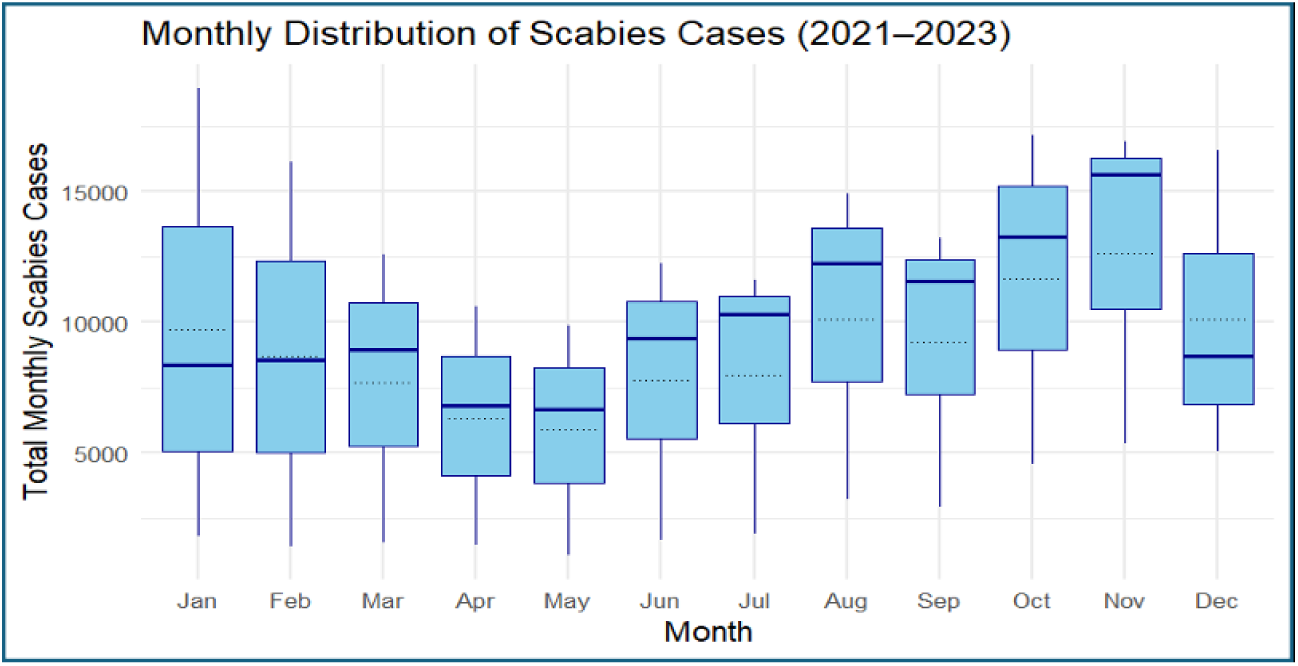
Monthly scabies case distribution in Rohingya camps (2021–2023)

April to May is the low season with lowest means and medians. The box heights are also narrow, indicating consistently lower cases in these months across the years.

June to September shows a gradual increase in median towards peak months.

To further assess and confirm seasonal pattern of scabies cases, a Seasonal-Trend decomposition using Loess (STL) was performed on monthly aggregated scabies case data from January 2021 to December 2023 as shown in Figure 3.

**Figure 3.**
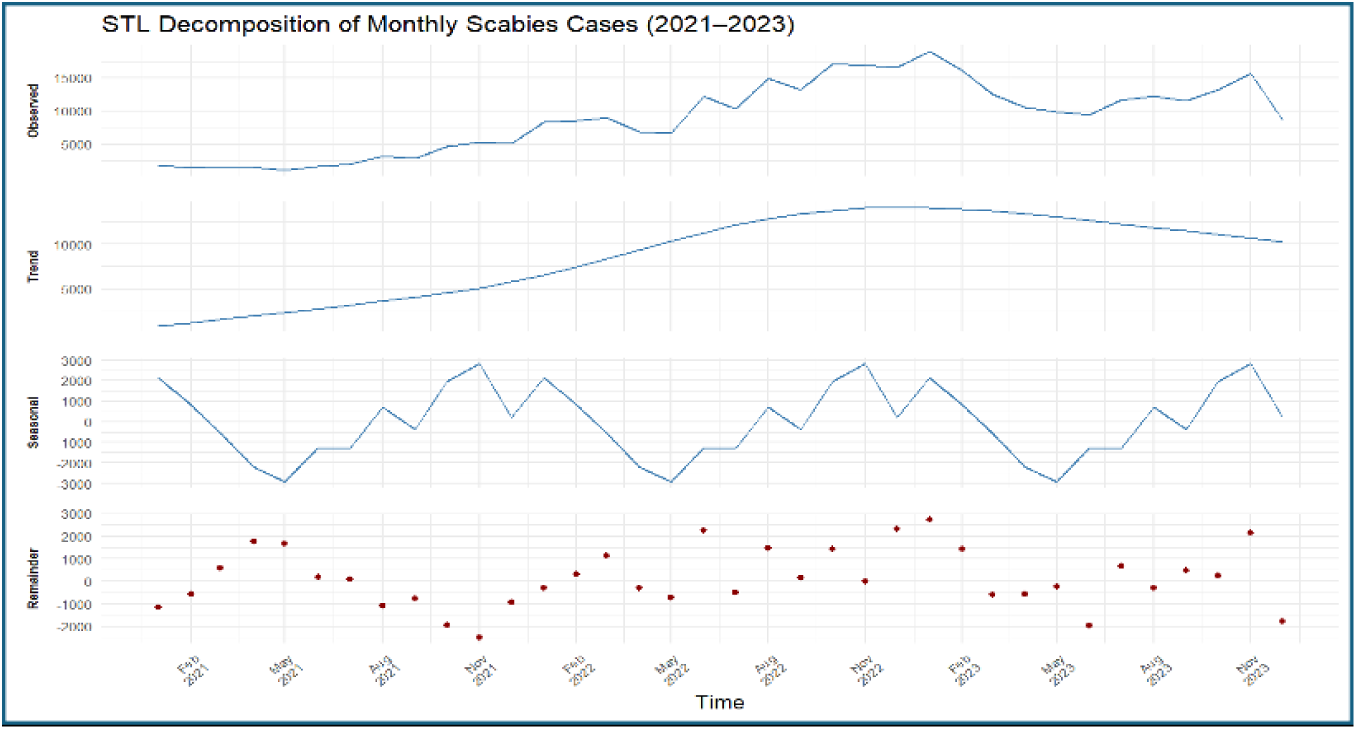
STL decomposition of monthly scabies cases in Rohingya camps (2021–2023)

The observed component demonstrates the observed pattern of scabies over the three-year period. The line indicates a gradual increase in scabies cases, with a peak around January-February 2023, followed by a decrease before rising again in October-November 2023. There was a sharp decline in December 2023, likely reflecting early programmatic interventions preceding formal mass drug administration.

The trend component shows a steady rise in the number of scabies cases from 2021 towards the peak in early 2023 followed by a slight decline, indicating a true rise of scabies over time irrespective of seasonality.

The third component, seasonality, demonstrates a clear and repeated seasonal pattern of scabies. Similar to the box plot, the increase of the scabies cases is seen from November to February (high-season) with the top peak in October to November; and a drop is in April to June (low-season).

The fourth and last component, remainder, shows random scattered distribution of cases indicating there are some random spikes unexplained by trend and seasonality.

Figure 4 demonstrates a scaled graph where climate variables are overlayed on the decomposed seasonal trend. All values (maximum, minimum and average temperature, humidity, rainfall and seasonal effect of scabies) are scaled between 0 and 1 to allow comparison on the same Y-axis. The graph shows an inverse relationship between temperature and number of scabies. Scabies cases increase from October to March when temperatures are lower and decrease during the warmer months of April to June.

**Figure 4.**
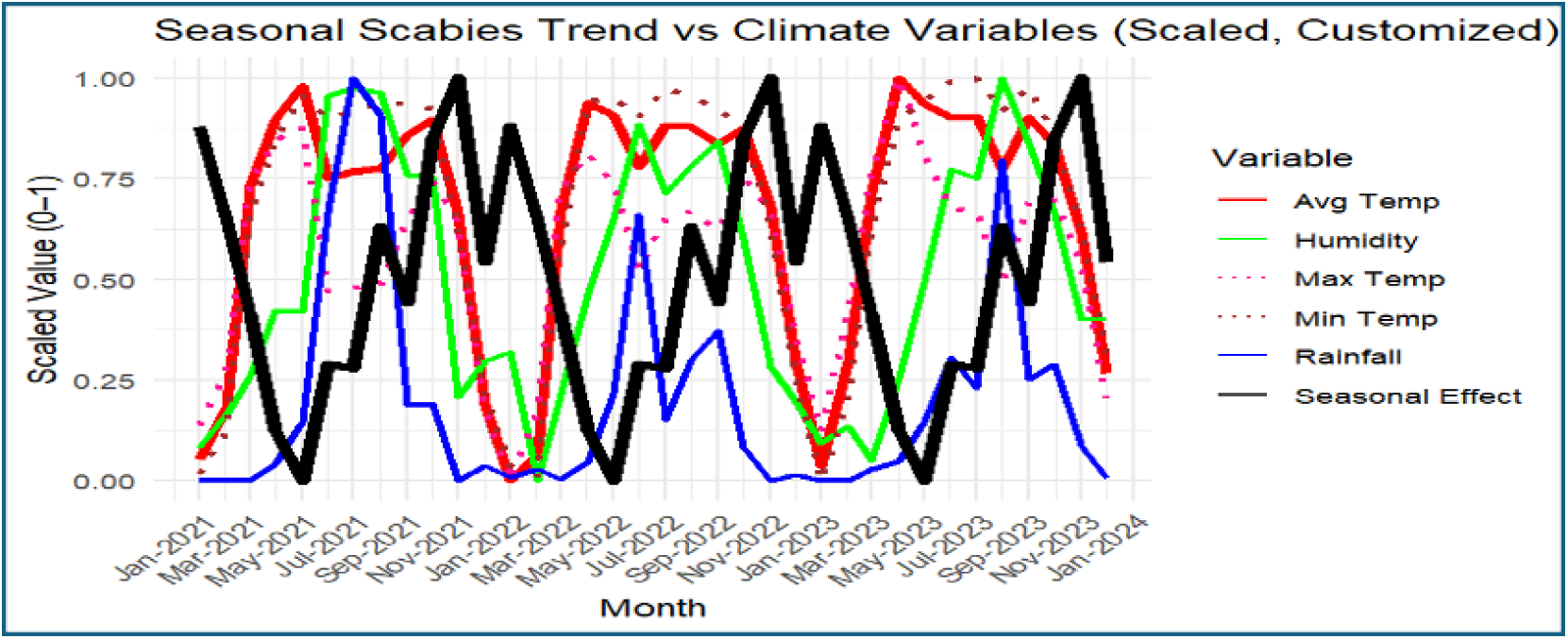
Seasonal scabies trend overlaid with climate variables (2021–2023)

No consistent relationship has been observed between the incidence of scabies cases and variations in rainfall or humidity.

A Pearson correlation test between climate variables and the decomposed seasonal component of scabies cases confirms the above findings (Table 3). It shows strong and significant negative correlation between seasonal rise and fall of scabies and all variables of temperature – maximum (r = -0.492, p = 0.002), minimum (r = -0.506, p = 0.002) and average (r = -0.525, p = 0.001). There is no significant correlation of the scabies seasonality with humidity or rainfall.

**Table 3.** Correlation between climate variables and scabies cases (overall and seasonal components), 2021–2023.

| Variable | Correlation with<br>overall scabies cases |  | Correlation with<br>decomposed seasonal<br>scabies cases |  |
| --- | --- | --- | --- | --- |
|  | r | p value | r | p value |
| Maximum Temperature | -0.014 | 0.937 | -0.492 | **0.002** |
| Minimum Temperature | -0.08 | 0.643 | -0.506 | **0.002** |
| Average Temperature | -0.053 | 0.758 | -0.525 | **0.001** |
| Humidity | -0.095 | 0.58 | -0.265 | 0.119 |
| Rainfall | -0.17 | 0.322 | -0.193 | 0.259 |

However, no significant correlation of climate variables was observed with the overall number of scabies cases, indicating influence of non-climatic factors on the overall number of scabies cases, beyond seasonality.

## DISCUSSION

Our study explored the seasonality of scabies and its association with climatic factors. As per the author’s knowledge, this is among the few studies in South Asia and first study in refugee context that explored the seasonality and climate correlation of scabies. The study found the evidence of seasonal pattern of scabies and found strong association of temperature on the seasonal trend of scabies. However, we found that scabies incidence is influenced by factors beyond seasonality, there should have strong influence of non-climatic factors.

The study found strong seasonality of scabies, where April to June is the low season corresponding to the summer and October to February is the high season corresponding to the cooler season (Later autumn, winter and early spring). Our study aligns with the findings of some other studies that explored seasonality of Scabies. For instance, a survey among school-age children in UK and a study of young adults in Israel found scabies infestation is higher in winter than summer. A study in Poland found highest number of scabies cases reported in Autumn and winter (20–22).

Seasonality of scabies can potentially be explained by climate factors and human behavior and social factors. April to June, when scabies case load was consistently found low, were having the highest temperature of the year. Our study also found that there is strong and significant negative correlation between seasonality of scabies and temperature indicating scabies case load drops with the rise of temperature. Similar correlation was also noted by a nation-wide study in Poland as well as a study in Taiwan (17,20). Studies found that mites can survive for prolonged period for 5-6 days in cold temperature (10–15 °C) and die within 1-2 days in high temperature (above 25 °C) (23). This could partially explain why the caseload was lower in summer and gradually went up towards peak in late autumn and winter.

There could also be some human behavior and social factors influencing seasonality of scabies. During winter, people have more tendency to crowd together inside the shelter and share and re-use bedding and clothing (e.g. blankets, warm clothes) without washing (24). In displacement setting, like Rohingya refugee camps, this is more pronounced due to poverty, insufficient provision of water, soap and sanitation, and fragile and climate resistant shelters (25–28). Conversely, in summer, people wear light clothes, which are easy to wash, and people avoid crowding and close contact due to extreme heat.

Although, we have found strong seasonal patterns of scabies and strong correlation between seasonal trend (decomposed) and temperature, the overall number of scabies are not significantly correlated with any climate variable. This explains that seasonality and climatic factors may partially explain seasonal variation in scabies trends and upsurge. All other non-climatic factors, including overcrowding, water, sanitation and hygiene, human behavior, public health interventions, MDA campaign and the availability of preventive and treatment services, have impactful influence on the overall incidence of scabies in the Rohingya refugee camps.

## LIMITATIONS OF THE STUDY

This study has several limitations. First, the analysis was based on routinely collected health facility surveillance data, which may be subject to underreporting, variation in healthcare-seeking behaviour, and differences in diagnostic practices across facilities. However, standardized clinical guidelines and routine data quality checks were applied across all IOM-supported facilities throughout the study period.

Second, the use of aggregated data precluded individual-level analysis and adjustment for potential confounders such as household crowding, water and sanitation conditions, or treatment adherence. As a result, the observed associations between scabies incidence and climatic variables should be interpreted as ecological relationships rather than causal effects.

Third, climate data were obtained from a central meteorological source and may not fully capture microclimatic variation within individual camps. Finally, data from 2024 onwards were excluded due to the implementation of mass drug administration, which substantially altered transmission dynamics and limited comparability with earlier years.

Despite these limitations, the large sample size, multi-year observation period, and consistent surveillance framework provide robust insight into seasonal patterns of scabies in a humanitarian setting.

## CONCLUSION

This study demonstrates clear seasonal variation in scabies incidence in the Rohingya refugee camps and surrounding host communities, with higher caseloads during late autumn and winter months and lower caseloads during warmer periods. Scabies incidence showed a consistent negative association with temperature, while no significant association was observed with rainfall or humidity.

These findings highlight the importance of incorporating seasonal patterns into scabies surveillance and preparedness planning in humanitarian settings. Timely implementation of public health measures, including intensified case management, targeted mass drug administration, and community-based prevention activities, may benefit from alignment with periods of increased risk. Further prospective and multi-site studies using individual-level data are needed to better understand underlying transmission dynamics and to inform context-specific control strategies.

## AUTHORS’ CONTRIBUTION

CEH conceptualized the study, supervised the analysis, and led manuscript development. MAH contributed to study design, data analysis, and drafting of the manuscript. JTP, HT, and MSH supported data collection and field coordination. MAH and MFH contributed to climate data acquisition and interpretation. ERS, JCO, UMP, and MFK critically reviewed the manuscript and provided technical input. All authors approved the final manuscript. CEH is the corresponding author and JCO is the guarantor.

## PATIENT CONSENT

Informed consent was not required. Investigators adhered to all data-security and confidentiality safeguards.

## DATA AVAILABILITY

All relevant aggregated data and analysis scripts are available from Zenodo: https://doi.org/10.5281/zenodo.17648621. Related datasets from the same scabies research series are available at: https://doi.org/10.5281/zenodo.17697474 and https://doi.org/10.5281/zenodo.17687717.

## Data Availability

All relevant aggregated data and analysis scripts generated during this study are available in the Zenodo repository at https://doi.org/10.5281/zenodo.17648621
. The data include anonymised, aggregated scabies case counts and corresponding climate variables for the period 2021 to 2023.

## ACKNOWLEDGMENTS

CEH conceptualized and designed the study, provided overall methodological guidance, and led the drafting and critical revision of the manuscript. MAH assisted in refining the study design and drafting the manuscript. JTP, HT supported field coordination, MSH assisted supporting the data collection processes. MAH supported with climatic data triangulation. ERS and JCO contributed to the critical review and refinement of the manuscript. MFK, ERS, UMP and JCO provided strategic oversight and ensured alignment of the study within broader programmatic and policy frameworks. All authors reviewed and approved the final version of the manuscript. CEH is the corresponding author and JCO is the guarantor of the study.

## GENERATIVE AI DISCLOSURE

Language editing assistance was provided using Grammarly (version 6.8.263) and ChatGPT (OpenAI; GPT-5 Thinking, accessed [November 2025]), limited strictly to grammar and clarity. No analyses, figures, or scientific content were generated using AI tools. All authors reviewed and took full responsibility for the content.

